# Household finished flooring and soil-transmitted helminth and *Giardia* infections among children in rural Bangladesh and Kenya: a prospective cohort study

**DOI:** 10.1101/2020.06.23.20138578

**Authors:** Jade Benjamin-Chung, Yoshika S. Crider, Andrew Mertens, Ayse Ercumen, Amy J. Pickering, Audrie Lin, Lauren Steinbaum, Jenna Swarthout, Mahbubur Rahman, Sarker M. Parvez, Rashidul Haque, Sammy M. Njenga, Jimmy Kihara, Clair Null, Stephen P. Luby, John M. Colford, Benjamin F. Arnold

**Affiliations:** Division of Epidemiology & Biostatistics, University of California, Berkeley, 2121 Berkeley Way West Rm 5302, Berkeley, CA, 94720 USA; Energy & Resources Group, University of California, Berkeley, 310 Barrows Hall, Berkeley, CA, 94720 USA; Department of Forestry and Environmental Resources, North Carolina State University, 2800 Faucette Dr, Raleigh, NC, 27607 USA; Civil and Environmental Engineering, Tufts University, 200 College Ave, Medford, MA, 02155 USA; Exponent, 980 9^th^ Street, 16^th^ Floor, Sacramento, CA, 95814 USA; International Centre for Diarrhoeal Disease Research, 68 Shaheed Tajuddin Ahmed Ave, Dhaka, 1212 Bangladesh; Eastern and Southern Africa Centre of International Parasite Control, Kenya Medical Research Institute, P.O. Box 54840 00200 Off Mbagathi Rd, Nairobi, Kenya; Center for International Policy Research and Evaluation, Mathematica Policy Research, 1100 First St NE #1200, Washington, DC 20002 USA; Division of Infectious Diseases and Geographic Medicine, Stanford University, 291 Campus Drive Li Ka Shing Building, Stanford, CA 94305 USA; Francis I. Proctor Foundation, University of California, San Francisco, 95 Kirkham St, San Francisco, CA 94122 USA; Department of Ophthalmology, University of California, San Francisco, 1426 Fillmore St, San Francisco, CA 94115

**Author notes:** Corresponding author: Jade Benjamin-Chung.

## Abstract

**Background:** Soil-transmitted helminths (STH) and *Giardia duodenalis* are responsible for a large burden of disease globally. In low-resource settings, household finished floors (e.g., concrete floors) may reduce transmission of STH and *G. duodenalis*.

**Methods:** In a prospective cohort of children nested within two randomised trials in rural Bangladesh and Kenya, we estimated associations between household finished flooring and STH and *G. duodenalis* prevalence. In 2015-2016, we collected stool samples from children aged 2-16 years in rural Bangladesh and Kenya. We detected STH infection using qPCR (Bangladesh N=2,800; Kenya N=3,094) and detected *G. duodenalis* using qPCR in Bangladesh (N=6,894) and ELISA in Kenya (N=8,899). We estimated adjusted prevalence ratios (aPRs) using log-linear models adjusted for potential confounders.

**Findings:** At enrolment, 10% of households in Bangladesh and 5% in Kenya had finished floors. In both countries, household finished flooring was associated with lower *Ascaris lumbricoides* prevalence (Bangladesh aPR: 0.33, 95% CI 0.14, 0.78; Kenya aPR: 0.62, 95% CI 0.39, 0.98) and any STH (Bangladesh aPR: 0.73, 95% CI 0.52, 1.01; Kenya aPR: 0.57, 95% CI 0.37, 0.88). Household finished floors were also associated with lower *Necator americanus* prevalence in Bangladesh (aPR: 0.52, 95% CI 0.29, 0.94) and *G. duodenalis* prevalence in both countries (Bangladesh aPR: 0.78, 95% CI 0.64, 0.95; Kenya: aPR: 0.82, 95% CI 0.70, 0.97).

**Interpretation:** In low-resource settings, living in households with finished floors over a two-year period was associated with lower prevalence of *G. duodenalis* and certain STH in children.

**Funding:** Bill & Melinda Gates Foundation grant OPPGD759

## Introduction

Soil-transmitted helminths (STH), including *Ascaris lumbricoides*, hookworm, and *Trichuris trichiura*, and *Giardia duodenalis* infect approximately 1.45 billion and 180 million people globally.^1,2^ Infections can result in anemia, malnutrition, and deficits in child physical and cognitive development.^3,4^ Mass preventive chemotherapy is the primary STH control strategy, however, its effectiveness is limited by frequent re-infection following deworming, low deworming efficacy against *T. trichiura*, and potential drug resistance.^5–8^ Improved water, sanitation, and hygiene (WASH) interventions may also reduce transmission because STH and *G. duodenalis* are transmitted through oral ingestion or dermal contact (in the case of hookworm) with fecally contaminated soil, food, water, or fomites.^9,10^ Recent trials of household WASH interventions reported moderate reductions in STH and *G. duodenalis* prevalence, but effect sizes varied between countries and interventions.^11–13^ A major challenge of household WASH interventions is that they require sustained behavior change and infrastructure maintenance to be effective over long periods of time.^14,15^

Housing improvements are associated with improved health.^16^ In particular, installation of household finished flooring (e.g., wood, cement, or tile) may interrupt STH and *G. duodenalis* transmission and does not require sustained behavior change or extensive maintenance typical of many WASH interventions. Finished flooring may decrease STH and *G. duodenalis* transmission by reducing fecal contamination of hands, food, and water. Microorganisms in household soil contribute importantly to hand contamination.^17^ Since STH ova survive best under moist conditions, impermeable finished flooring may reduce environmental survival and interrupt the life cycle of STH, which requires a soil stage for STH ova to become infective.^3,18^ Studies in rural Kenya and urban Brazil have detected STH ova inside households at concentrations comparable to samples collected near latrines.^19,20^ Structured observations in low-resource settings have found that young children frequently ingest soil in households with dirt floors^21^ and that the presence of STH larvae in soil near child play areas is associated with increased STH infection.^22^ These studies support the biological plausibility of household finished flooring as an environmental intervention to reduce enteric parasite transmission.

Cross-sectional studies in urban and rural settings have reported associations between living in a household with a soil floor and higher risk of infection with hookworm^23^, *A. lumbricoides*^22,24,25^, *Strongyloides stercoralis*^26^, *G. duodenalis*^24,27^, cholera,^28^ other enteric parasite infections,^29–31^ and diarrhea or gastrointestinal illness.^32,33^ A retrospective, matched impact evaluation of a government program in urban Mexico found that replacing household dirt floors with concrete reduced diarrhea and intestinal parasite infection in children.^34^ However, this study did not describe the parasite species that were measured.

Here, we report the findings of a prospective cohort study of children nested within two cluster-randomised trials in rural Bangladesh and Kenya. We investigated whether young children living in homes with finished floors (wood, tile, concrete) had lower prevalence and intensity of STH and *G. duodenalis* infection two years after enrolment compared to children living in homes with unfinished (earthen) floors. Our study improves upon prior studies by using prospectively collected data and molecular detection methods.^35^ In addition, study populations included a cohort born into households with different flooring material, allowing us to investigate associations between household flooring and infection in early life.

## Methods

### Study design

We conducted a prospective, observational analysis of children enrolled in the WASH Benefits cluster-randomised trials in Bangladesh and Kenya. The trials delivered individual and combined water, sanitation, handwashing, and nutrition interventions to clusters of households to estimate impacts on child growth, diarrhea, and enteric infection 2 years after enrolment. Study arms included improved water (W), improved sanitation (S), improved handwashing (H), combined W+S+H, breastfeeding promotion and lipid-based nutrient supplements (N), combined W+S+H+N, and a double-sized control arm. In this analysis, we pooled across the study arms in each trial. The Bangladesh trial was conducted in rural villages in Gazipur, Kishoreganj, Mymensingh, and Tangail districts, and the Kenya trial was conducted in rural villages in western Kenya in Bungoma, Kakamega, and Vihiga counties. Additional trial details have been reported elsewhere.^11–13,36,37^

### Participants

Both trials defined birth cohorts by enrolling pregnant women and following their live births until approximately age 2 years. Children aged 18-27 months were also enrolled if a child of that age was present in the household. Enrolment occurred from May 2012 and July 2013 in Bangladesh and between November 2012 and May 2014 in Kenya. At approximately 2 years follow-up, the trials enrolled additional school-aged children in the same households or compounds as children in the birth cohort. The Bangladesh trial enrolled up to 2 other children aged 3-12 years per compound. The Kenya trial enrolled one school-aged child per household by selecting the youngest available child aged 3-15 years. These additional enrolments occurred at the time of stool sample collection (May 2015-May 2016 in Bangladesh and January 2015-July 2016 in Kenya).

### Exposure measurement

Field staff observed household flooring material at enrolment and two-year follow-up. We classified household floors observed at enrolment as “finished” if the predominant floor material was wood, tile, or concrete and as “unfinished” if the floor material was partially made of wood, tile, or concrete or entirely made of soil.

### Outcome measurement

We pre-specified the following primary outcomes: prevalence of any STH infection measured by quantitative polymerase chain reaction (qPCR) and prevalence of *G. duodenalis* infection measured by qPCR in Bangladesh and ELISA in Kenya. Secondary outcomes included species-specific prevalence and mean cycle quantification (Cq) values using qPCR and prevalence, mean eggs per gram, and moderate/heavy infection intensity measured by Kato-Katz. Studies tested for STH with multi-parallel qPCR in a subsample of study arms (control, S, W, W+S+H in Bangladesh; control, W+S+H, and W+S+H+N in Kenya), and with double-slide Kato-Katz in all study arms. We chose to focus our primary analysis on qPCR outcomes because it has higher sensitivity and specificity than Kato-Katz in settings with predominantly low infection intensity and because of potential misclassification of *A. lumbricoides* using Kato-Katz in the Bangladesh trial.^38,39^ We categorized STH infection intensity using Kato-Katz as low, moderate, or heavy for hookworm (1-4999, 5000-49999, 50000+ eggs per gram (epg)), *A. lumbricoides* (1-4999, 5000-49999, 50000+ epg), and *T. trichiura* (1-999, 1000-9999, 10000+ epg).^40^ Additional details on outcome measurement are in Appendix 1 and are available elsewhere.^11–13,38^

### Study size

We estimated the minimum detectable effect treating the sample size in all study arms in each country as fixed and assuming 80% statistical power and a type I error rate of 5%. We calculated prevalence of any STH and *G. duodenalis* and the design effect accounting for correlation at the cluster level in each country. Prevalence of any STH was 36% in Bangladesh and 29% in Kenya; *G. duodenalis* prevalence was 33% in Bangladesh and 39% in Kenya. The design effect was for any STH prevalence was 1.9 in Bangladesh and 1.7 in Kenya; for *G. duodenalis* prevalence it was 1.5 in Bangladesh and 1.2 in Kenya. Minimum detectable prevalence ratios for any STH were 0.67 in Bangladesh and 0.58 in Kenya; for *G. duodenalis*, they were 0.60 in Bangladesh and 0.82 in Kenya.

### Statistical methods

We pre-specified the analysis plan for this study at https://osf.io/mtf8x/. We conducted analyses separately by country. We estimated prevalence ratios and prevalence differences using log-linear modified Poisson models.^41^ We estimated the relative reduction in mean Cq values (the ratio of the arithmetic mean of Cq values – 1) among children who were infected using linear models and used the delta method to obtain standard errors. We also estimated associations using double-robust, targeted maximum likelihood estimation (TMLE) to optimize bias-variance tradeoff.^42^ Adjusted models controlled for each of the following pre-specified covariates that were associated with the outcome (likelihood ratio test p-values < 0.2): month of sample collection, child age, child sex, child birth order (Bangladesh only), mother’s age, mother’s educational attainment (no education, primary, secondary), mother’s height (cm), number of individuals in the household ≤ 18 years of age, the number of individuals in compound, food insecurity (using the Household Food Insecurity Access Scale^43^), household electricity, and household assets (e.g., table or tv ownership). We also included study arm as a covariate to adjust for any effects of interventions. We did not adjust for categorical covariates with prevalence < 5%. Robust standard errors were clustered at the village cluster level.^44^ We assessed potential effect modification (pre-specified) by birth cohort enrolment (vs. older children), improved latrine access at enrolment, and caregiver reported deworming in last 6 months.

To diagnose and prevent positivity violations (a lack of experimentation in the exposure in a covariate stratum),^45^ we used an ensemble machine learning algorithm^46^ to estimate the predicted probability of having a finished floor adjusting for covariates defined above. As a sensitivity analysis, we excluded observations with extreme predicted probabilities of having finished flooring. We excluded observations within 1% of the minimum and maximum predicted probabilities in each country. A small proportion of households changed flooring status between enrolment and two-year follow-up. We conducted a sensitivity analysis excluding households that changed status and a separate sensitivity analysis classifying households that improved their flooring status during the follow-up period as having a finished floor.

To assess the influence of confounding on our estimates, we estimated E-values, which quantify the magnitude of association that an unmeasured confounder would be required to have with the exposure and each outcome for the unmeasured confounder to be completely responsible for a given result.^47^ Large E-values indicate that an unmeasured confounder would need to have strong associations with the exposure and outcome to fully explain our findings, whereas small E-values indicate that unmeasured confounding may be a larger concern for a given estimate.

Replication scripts and data are available at https://osf.io/dgkw5/, and a permanent archive of the code associated with this preprint is available at https://github.com/jadebc/washb-floors-public/releases/tag/medRxiv. This study was performed using publicly available, de-identified data and was exempt from human subjects committee approval. The original trials were approved by Committee for the Protection of Human Subjects at the University of California, Berkeley (2011-09-3652, 2011-09-3654), the Institutional Review Board at Stanford University (23310, 25863), the Scientific and Ethics Review Unit at the Kenya Medical Research Institute (SSC-2271), and the Ethical Review Committee at The International Centre for Diarrhoeal Disease Research, Bangladesh (PR-11063).

## Results

In Bangladesh, from 7,795 children enrolled at two-year follow-up, 7,187 provided stool specimens (Appendix Figure 1). In Kenya, from 9,686 children enrolled at two-year follow-up, 9,077 provided stool specimens. Kato-Katz was used to detect STH in 100% of stool specimens in Bangladesh and 99.6% of specimens in Kenya. qPCR was used to detect STH in a subsample in each country (Bangladesh N = 2,800; Kenya N = 3,093). Sufficient stool for an additional aliquot was available to detect *G. duodenalis* in 6,894 specimens in Bangladesh and 8,899 specimens in Kenya. In Bangladesh, the mean child age was 4.8 years (range: 1.8-12 years). In Kenya the mean child age was 3.6 years (range: 0.2-16 years). 65% of children in Bangladesh and 42% of children in Kenya were reported to have consumed deworming medication in the previous 6 months.

The prevalence of household finished flooring at enrolment was 10% in Bangladesh and 5% in Kenya. Fewer than 5% of households changed household flooring status between baseline and two-year follow-up. In each country, some maternal and household characteristics differed among households with and without household finished flooring at enrolment (Table 1). In Bangladesh, the proportion of mothers with at least some secondary education was 82.5% vs. 48.3% in households with vs. without finished floors. In both countries, households with finished floors were more likely to have electricity, improved wall and roof materials, and a television, motorcycle, and mobile phone compared to households without finished floors.

**Table 1.**
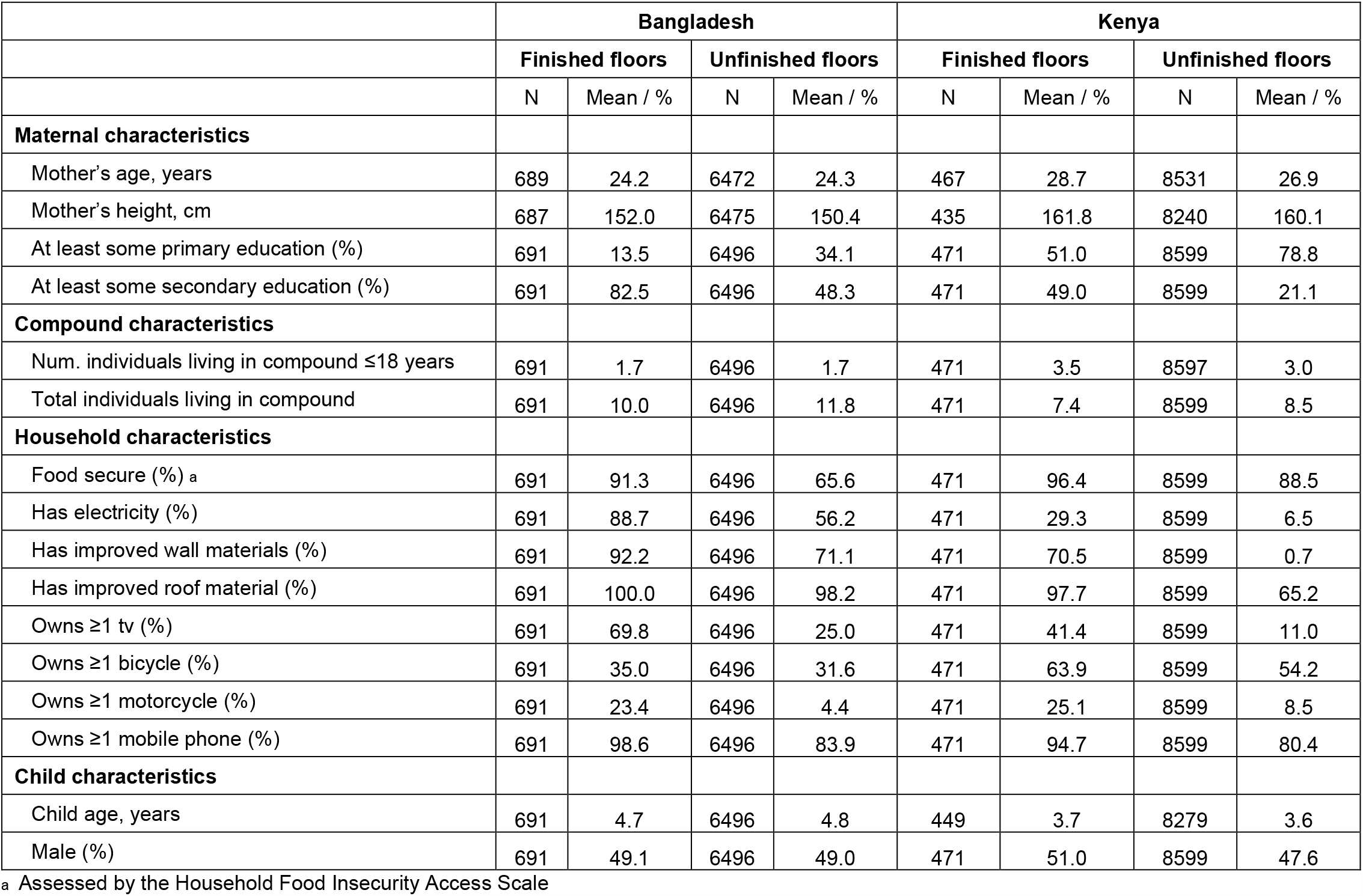
Characteristics at enrolment by household finished flooring status.

|  | Bangladesh |  |  |  | Kenya |  |  |  |
| --- | --- | --- | --- | --- | --- | --- | --- | --- |
|  | Finished floors |  | Unfinished floors |  | Finished floors |  | Unfinished floors |  |
|  | N | Mean / % | N | Mean / % | N | Mean / % | N | Mean / % |
| **Maternal characteristics** |  |  |  |  |  |  |  |  |
| Mother's age, years | 689 | 24.2 | 6472 | 24.3 | 467 | 28.7 | 8531 | 26.9 |
| Mother's height, cm | 687 | 152.0 | 6475 | 150.4 | 435 | 161.8 | 8240 | 160.1 |
| At least some primary education (%) | 691 | 13.5 | 6496 | 34.1 | 471 | 51.0 | 8599 | 78.8 |
| At least some secondary education (%) | 691 | 82.5 | 6496 | 48.3 | 471 | 49.0 | 8599 | 21.1 |
| **Compound characteristics** |  |  |  |  |  |  |  |  |
| Num. individuals living in compound ≤18 years | 691 | 1.7 | 6496 | 1.7 | 471 | 3.5 | 8597 | 3.0 |
| Total individuals living in compound | 691 | 10.0 | 6496 | 11.8 | 471 | 7.4 | 8599 | 8.5 |
| **Household characteristics** |  |  |  |  |  |  |  |  |
| Food secure (%) <sup>a</sup> | 691 | 91.3 | 6496 | 65.6 | 471 | 96.4 | 8599 | 88.5 |
| Has electricity (%) | 691 | 88.7 | 6496 | 56.2 | 471 | 29.3 | 8599 | 6.5 |
| Has improved wall materials (%) | 691 | 92.2 | 6496 | 71.1 | 471 | 70.5 | 8599 | 0.7 |
| Has improved roof material (%) | 691 | 100.0 | 6496 | 98.2 | 471 | 97.7 | 8599 | 65.2 |
| Owns ≥1 tv (%) | 691 | 69.8 | 6496 | 25.0 | 471 | 41.4 | 8599 | 11.0 |
| Owns ≥1 bicycle (%) | 691 | 35.0 | 6496 | 31.6 | 471 | 63.9 | 8599 | 54.2 |
| Owns ≥1 motorcycle (%) | 691 | 23.4 | 6496 | 4.4 | 471 | 25.1 | 8599 | 8.5 |
| Owns ≥1 mobile phone (%) | 691 | 98.6 | 6496 | 83.9 | 471 | 94.7 | 8599 | 80.4 |
| **Child characteristics** |  |  |  |  |  |  |  |  |
| Child age, years | 691 | 4.7 | 6496 | 4.8 | 449 | 3.7 | 8279 | 3.6 |
| Male (%) | 691 | 49.1 | 6496 | 49.0 | 471 | 51.0 | 8599 | 47.6 |

<sup>a</sup> Assessed by the Household Food Insecurity Access Scale

The prevalence of any STH infection using qPCR was 35% in Bangladesh and 28% in Kenya, and the prevalence of *G. duodenalis* was 32% in Bangladesh and 39% in Kenya. The percentage of children with moderate or heavy infection intensity using Kato-Katz was 4% for *A. lumbricoides* in Bangladesh and 12% in Kenya and <1% for hookworm and *T. trichiura* in both countries. Comparing households with finished vs. unfinished flooring, the prevalence of any STH infection was 14.5% vs. 36.7% in Bangladesh and 14.5% vs. 29.2% in Kenya using qPCR (Appendix Table 1). STH infection prevalence was substantially lower using Kato-Katz compared to qPCR (Appendix Table 2).

Unadjusted prevalence ratios (PR) for STH detected with qPCR indicated a protective association between household finished flooring and *A. lumbricoides, N. americanus, T. trichiura*, and any STH infection in Bangladesh and *A. lumbricoides* and any STH infection in Kenya (Figure 1). After adjusting for potential confounders, household finished flooring was associated with lower prevalence of *A. lumbricoides* in both countries (Bangladesh adjusted PR (aPR) = 0.34, 95% CI 0.15, 0.80; Kenya aPR = 0.61, 95% CI 0.39, 0.97) and *N. americanus* in Bangladesh (aPR = 0.52; 95% CI 0.29, 0.94). Associations between household finished flooring status and *A. ceylanicum* and *T. trichiura* prevalence in Bangladesh and *N. americanus* and *T. trichiura* in Kenya were null. For any STH prevalence, adjusted PRs were 0.73 (95% CI 0.51, 1.01) in Bangladesh and 0.57 (95% CI 0.37, 0.88) in Kenya. For STH detected using Kato-Katz, aPRs were similar to those detected by qPCR except that there was a statistically significant reduction in *T. trichiura* infection in Bangladesh using Kato-Katz that was not observed when using qPCR (aPR = 0.57, 95% CI 0.33, 0.96) (Appendix Table 3). Household finished floors were not associated with moderate/heavy *A. lumbricoides* infection in either country (Bangladesh aPR = 0.56, 95% CI 0.24, 1.31; Kenya aPR = 0.71, 95% CI 0.45, 1.10). In both countries, household finished flooring was associated with lower *G. duodenalis* prevalence (Bangladesh aPR = 0.78, 95% CI 0.64, 0.95; Kenya aPR = 0.82, 95% CI 0.70, 0.97) (Figure 1).

**Figure 1.**
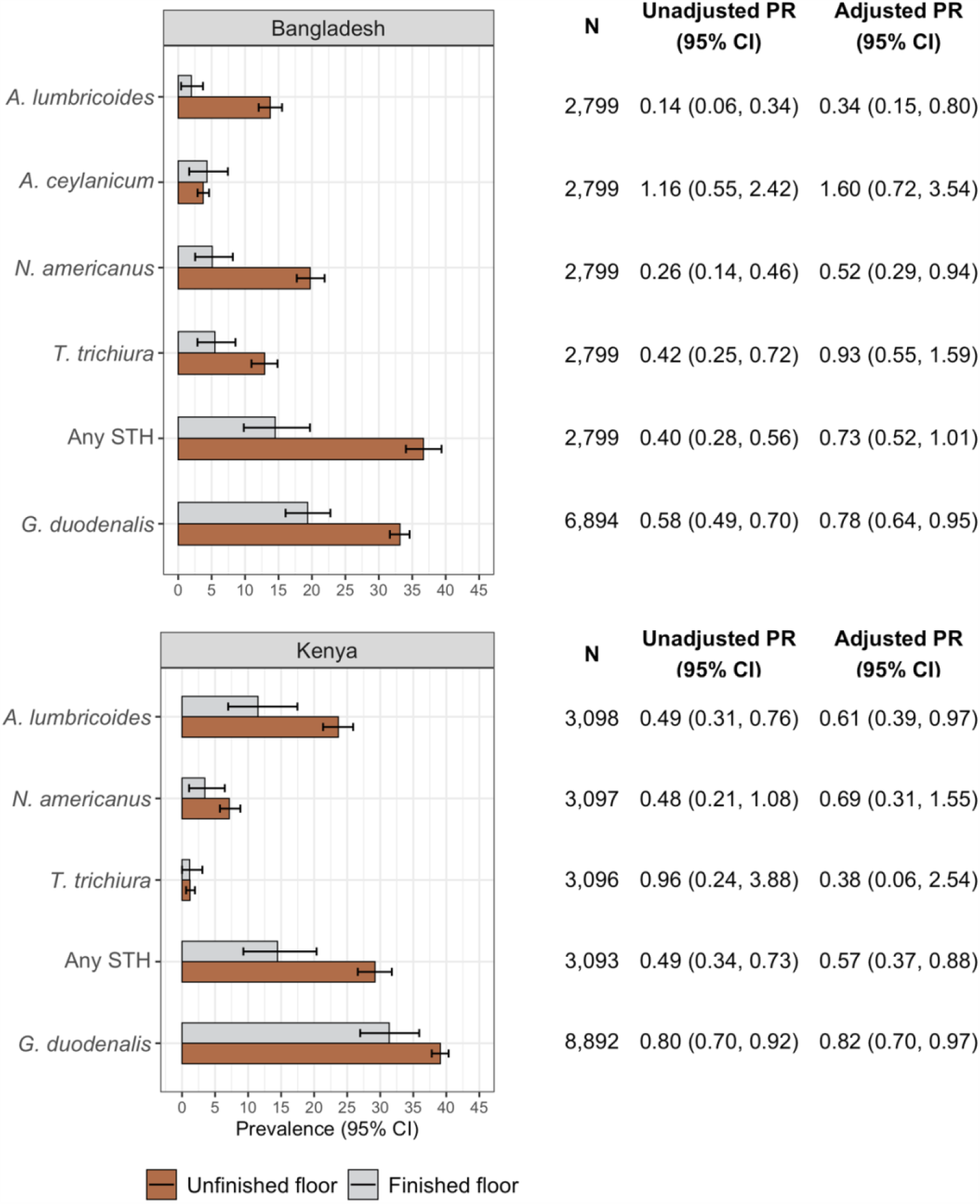
Soil-transmitted helminth and *Giardia duodenalis* prevalence and unadjusted and adjusted prevalence ratios comparing infection at two-year follow-up by household flooring status at enrolment. Finished floors includes wood, concrete, or tile household floors; unfinished floors were made of soil or earth. In the plots, error bars indicate 95% confidence intervals. Prevalence ratios (PRs) compared the prevalence of infection at two-year follow-up in children with improved household flooring at enrolment to those with unimproved household flooring at enrolment. Adjusted PRs control for potential confounders associated with the outcome (see list in the Methods section). Confidence intervals were adjusted for clustering at the village level. *Ancylostoma duodenale* was only detected in one sample in each country, so it is excluded from this figure.

In both countries, among children with infections detected by qPCR, mean Cq values were similar in children with vs. without finished flooring (Figure 2). Among children with STH infections detected by Kato-Katz, there were statistically significant adjusted fecal egg count reductions for *T. trichiura* in Bangladesh (−0.45, 95% CI −0.59, −0.32) (Appendix Table 4; Appendix Figure 2). Analyses using TMLE produced slightly different point estimates but overall were consistent with analyses using log-linear models (Appendix Table 5; Appendix Table 6).

**Figure 2.**
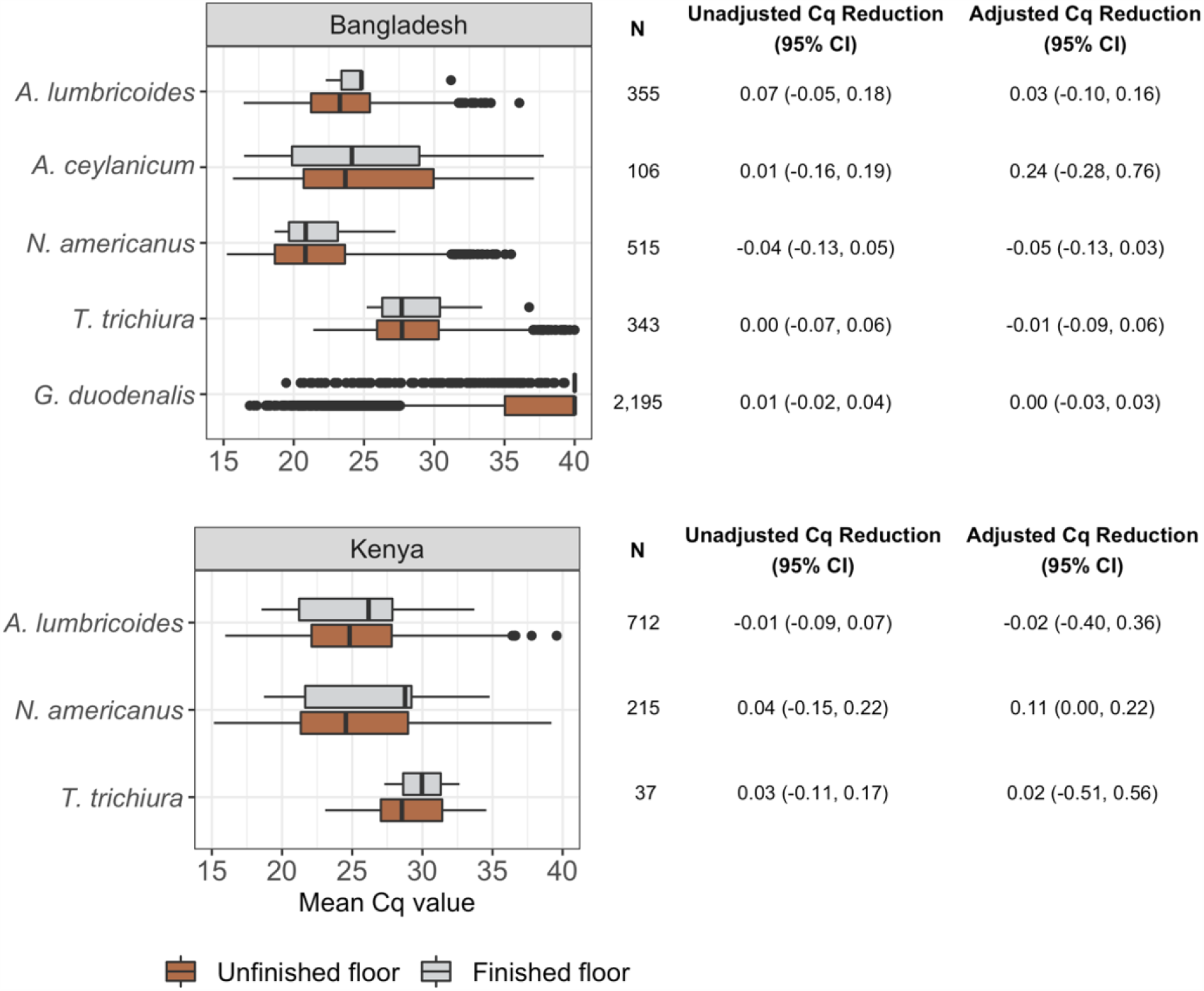
Soil-transmitted helminth and *Giardia duodenalis* mean Cq value and unadjusted and adjusted Cq reductions among infected individuals at two-year follow-up by household flooring status at enrolment. Finished floors includes wood, concrete, or tile household floors; unfinished floors were made of soil or earth. In the plots, error bars indicate 95% confidence intervals. Cq reductions were estimated as the ratio of the arithmetic mean Cq value at two-year follow-up in children with improved household flooring at enrolment to those with unimproved household flooring at enrolment minus 1. Adjusted Cq ratios control for potential confounders associated with the outcome (see list in the Methods section). Confidence intervals were adjusted for clustering at the village level. *Ancylostoma duodenale* was only detected in one sample in each country, so it is excluded from this figure.

There was no evidence of statistically significant effect modification on the multiplicative scale by child deworming consumption or improved latrine access at enrolment in either country (Appendix Figure 3). Compared to older children, children in the birth cohort had a stronger protective association between finished flooring and *G. duodenalis* prevalence in Bangladesh (birth cohort aPR = 0.68 (95% CI 0.52, 0.91); older children aPR = 0.86 (95% CI 0.69, 1.08); interaction p-value = 0.020).

In a sensitivity analysis excluding observations with extreme predicted probabilities of having a finished floor (Appendix Figure 4), in Kenya, household characteristics were very similar to the full set of observations (Appendix Table 7). In Bangladesh, household characteristics were similar in households with a finished floor; among those without a finished floor, there were lower levels of household electricity, food security, and other indicators of socioeconomic status after excluding observations with extreme predicted probabilities. Overall, adjusted prevalence ratios were similar when excluding observations with extreme predicted probabilities (Appendix Table 8). Sensitivity analyses reclassifying household flooring status to account for the small number of households that changed status during follow-up produced similar results to the primary analysis (Appendix Figure 5).

For outcomes with statistically significant associations in the primary analysis, the E-values ranged from 1.19 for *G. duodenalis* to 5.28 for *A. lumbricoides* in Bangladesh and from 1.43 for *G. duodenalis* to 1.87 for *A. lumbricoides* in Kenya (Appendix Table 9).

## Discussion

We used a prospective, observational design to estimate associations between household finished flooring and STH and *G. duodenalis* infection in young children. In Bangladesh, we found protective associations between household finished flooring and prevalence of *A. lumbricoides, N. americanus*, any STH, and *G. duodenalis* but not *A. ceylanicum and T. trichiura*. In Kenya, we found protective associations with *A. lumbricoides*, any STH, and *G. duodenalis* but not *N. americanus* or *T. trichiura*. Among infected individuals, household finished flooring was not associated with lower mean Cq values but was associated with lower mean eggs per gram using Kato-Katz for *T. trichiura* in Bangladesh and *A. lumbricoides* in Kenya. While results varied by species of STH and by country, these results suggest that household finished floors hold promise as an environmental intervention to reduce transmission of STH and *G. duodenalis* among children in low-resource settings.

Our findings are consistent with the small number of prior studies that have used observational designs to estimate the association between household finished flooring and enteric illness. A cross-sectional study in Bangladesh reported significant protective associations of similar magnitude with *A. lumbricoides* and hookworm prevalence and also found no association with *T. trichiura* in children and women aged 15-49 years.^25^ A retrospective, matched study of a concrete flooring program in Mexico reported statistically significant 21% percent reductions in parasite count (species were not identified) and a statistically significant 13% percent reduction in diarrhea in children among households whose dirt floors were fully replaced by concrete floors.^34^ A cross-sectional study in Zimbabwe reported a statistically significant 18% percent lower diarrhea prevalence in infants living in homes with finished floors.^32^

The magnitude of protective associations for household flooring was similar to or greater than the typical effect size for WASH and preventive chemotherapy interventions. Recent trials reported that WASH interventions reduced STH prevalence by up to 33% (results varied by species and intervention)^11,12^ and *G. duodenalis* prevalence by up to 25% in Bangladesh^13^ (in Kenya interventions did not reduce prevalence^11^). A meta-analysis of randomised and quasi-randomised studies estimated that mass deworming for STH delivered for at least 6 months reduced STH prevalence by 77% for *A. lumbricoides* and 33% for hookworm (there was no effect on *T. trichiura*).^48^ In this study, household finished flooring was associated with 38-67% lower *A. lumbricoides* prevalence and 18-22% lower *G. duodenalis* prevalence in both countries and 48% lower *N. americanus* prevalence in Bangladesh. In prior studies, neither WASH interventions, preventive chemotherapy, nor household finished flooring have been observed to be associated with lower *T. trichiura* prevalence.^11,12,25,48^ Here, when using Kato-Katz, we found a significant 43% lower *T. trichiura* prevalence among children in households with finished floors.

Household finished floors have several advantages over other interventions to reduce STH and *G. duodenalis* infection. Compared to WASH interventions, household finished floors require minimal sustained behavior change and are easy to maintain. For control of STH infections, household finished flooring may complement preventive chemotherapy, which has high reinfection rates following treatment.^6^ Another potential advantage of flooring is increased happiness; while we did not measure this outcome in this study, a study in Mexico reported increased happiness of adult residents of homes with newly installed concrete floors.^34^

Household socioeconomic status is likely a strong confounder of household flooring material and enteric parasite infection, so it is possible that residual, unmeasured confounding influenced our findings.^49^ We assessed the role of unmeasured confounding using E-values,^47^ which suggest that for unmeasured confounding to fully explain our statistically significant findings, the relative-scale association between unmeasured confounders and our exposures and outcomes would need to be 1.19-5.28 in Bangladesh and 1.43-1.87 in Kenya. The impact of unmeasured confounding may be even stronger given that our exposure was relatively rare (10% of participants in Bangladesh, and 5% in Kenya). Future randomised studies of household flooring would be able to minimize unmeasured confounding to rigorously estimate the effect of household flooring on enteric parasite infection.

This study is subject to additional limitations. First, we only measured outcomes in children, but household finished flooring may also reduce infection among adults. We would expect that reductions would be largest among individuals who spend more time in the home, namely young children, women, and the elderly. Second, some STH species were rare in our study populations (e.g., *A. ceylanicum* in Bangladesh, hookworm species and *T. trichiura* in Kenya), so some estimates may have had low statistical power. Finally, a prior study found evidence of an interaction between household flooring and household WASH infrastructure for certain STH;^25^ while we investigated associations in these subgroups in this study, subgroup analyses may have been underpowered.

Strengths of this study include the use of rigorous molecular diagnostic methods to measure outcomes and prospective data collection. In addition, the inclusion of data from two countries with differing ecology and disease transmission dynamics increases the generalizability of this study to other rural low-resource settings. A large proportion of children in this study were enrolled at birth, allowing us to measure the association between infection and household flooring exposures from birth through early life. Our finding that associations for *G. duodenalis* were more strongly protective in pre-school aged children supports the biological plausibility of dirt floors as a source of exposure to fecal contamination in early life since younger children are most likely to be exposed in the home.

## Conclusion

Household finished flooring holds promise as a potential intervention to reduce enteric parasites such as STH and *G. duodenalis* in low-resource settings. Compared to WASH interventions, which also target enteric parasite transmission pathways, a key potential advantage of household finished flooring is that it requires minimal maintenance and behavior change. Our findings motivate future, randomised studies to rigorously assess the impact of household finished flooring on infection.

## Data Availability

Replication scripts and data are available at https://osf.io/dgkw5/, and a permanent archive of the code associated with this preprint is available at https://github.com/jadebc/washb-floors-public/releases/tag/medRxiv.

## Acknowledgements

The authors would like to acknowledge Anmol Seth and Stephanie Djajadi for their contributions to figure and table generation.

## Author contributions

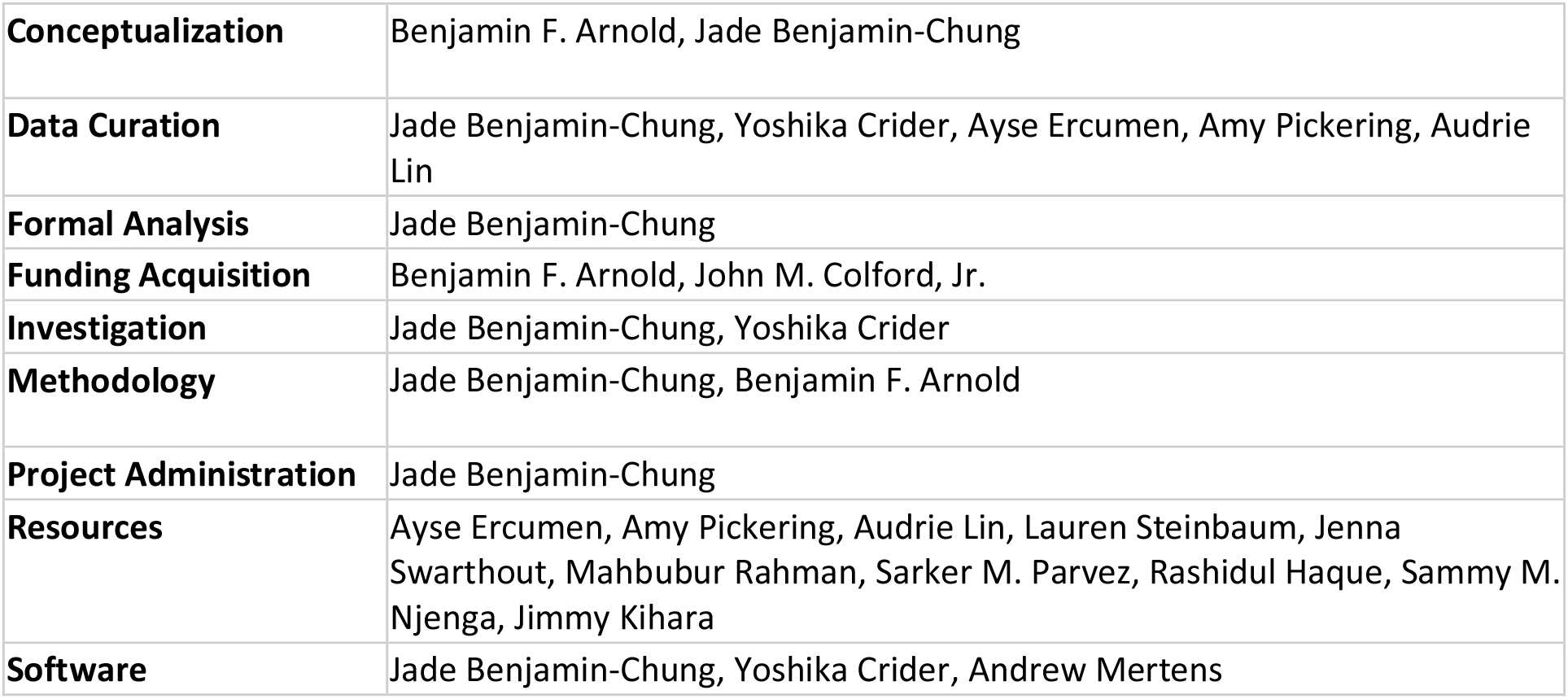
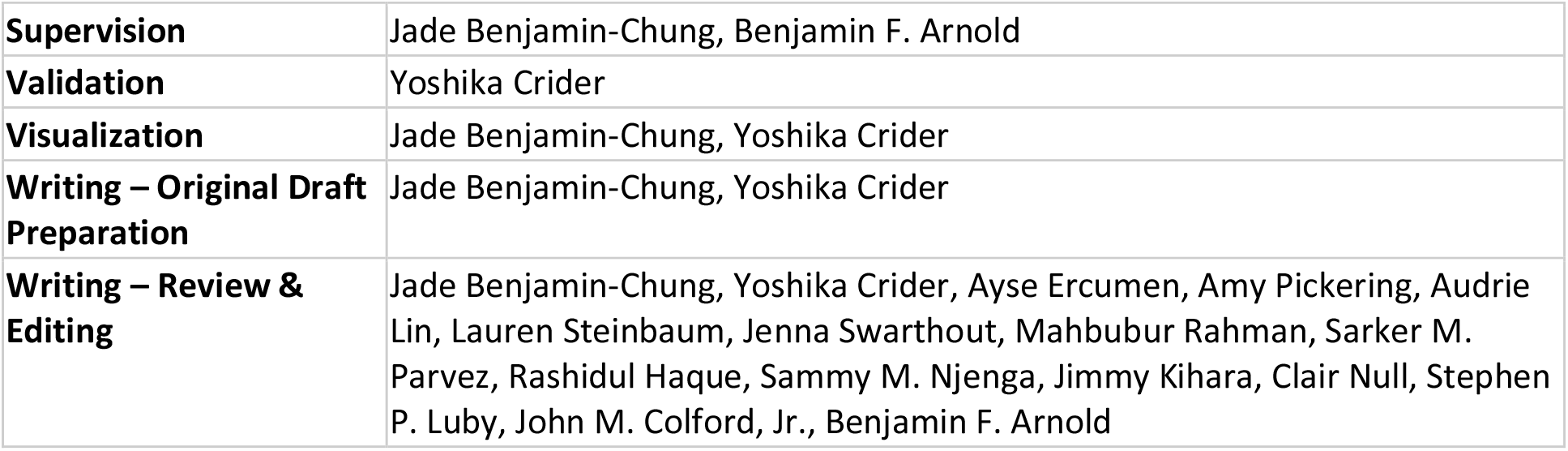

